# Self-Reported SARS-CoV-2 Vaccination is Consistent with Electronic Health Record Data among the COVID-19 Community Research Partnership

**DOI:** 10.1101/2022.05.13.22275054

**Authors:** Ashley H. Tjaden, Lida M. Fete, Sharon L. Edelstein, Michael Gibbs, Amy Hinkelman, Michael Runyon, Roberto P. Santos, William S. Weintraub, Joshua Yukich, Diane Uschner, the COVID-19 Community Research Partnership Study Group

## Abstract

**Introduction:** Observational studies of SARS-CoV-2 vaccine effectiveness depend on accurate ascertainment of vaccination receipt, date, and product type. Self-reported vaccine data may be more readily available to and less expensive for researchers than assessing medical records.

**Methods:** We surveyed adult participants in the COVID-19 Community Research Partnership who had an authenticated EHR (N=41,484) concerning receipt of SARS-CoV-2 vaccination using a daily survey beginning in December 2020 and a supplemental survey in September-October 2021. We compared self-reported information to that available in the EHR for the following data points: vaccine brand, date of first dose, and number of doses. Self-reported data was available immediately following vaccination (in the daily survey) and at a delayed interval (in the supplemental survey).

**Results:** For the date of first vaccine dose, self-reported “immediate” recall was within +/- 7 days of the date reported in the “delayed” survey for 87.9% of participants. Among the 19.6% of participants with evidence of vaccination in their EHR, 95% self-reported vaccination in one of the two surveys. Self-reported dates were within +/- 7 days of documented EHR vaccination for 97.6% of the “immediate” surveys and 92.0% of the “delayed” surveys. Self-reported vaccine product details matched those in the EHR for over 98% of participants for both “immediate” and “delayed” surveys.

**Conclusions:** Self-reported dates and product details for COVID-19 vaccination can be a good surrogate when medical records are unavailable in large observational studies. A secondary confirmation of dates for a subset of participants with EHR data will provide internal validity.

## INTRODUCTION

Observational studies of SARS-CoV-2 vaccine effectiveness (VE) and time to seroconversion or seroreversion depend on accurate ascertainment of vaccination receipt, date, and product type. While medical records of vaccination are typically considered the gold standard, self-reported vaccine data may be more readily available and less expensive.

In their guidance on how to assess COVID-19 VE using observational studies, the WHO advised against the use of self-reported COVID-19 vaccination data as the sole source indicating whether a person is vaccinated for primary analyses, due to recall bias and lack of product details.^1^ Self-reported dates are subject to recall bias, but Electronic Health Record (EHR) data may contain vaccination status only on a subset of participants and may also be erroneous.^2,3^ In the United States, COVID-19 vaccines have been made available in both medical and non-medical settings, including hospitals, pharmacies, public health departments, and mass vaccination sites, and the information may not be reliably recorded in a participant’s EHR; thus, self-reported data may be the only source for obtaining real-time vaccination information in a disease surveillance study.

Researchers have assessed the validity of self-reported influenza vaccine status in various populations and have found self-reported influenza vaccine status to be a very sensitive and moderately specific indicator of actual vaccine status.^4,5^ Researchers have found slightly lower, yet still high, sensitivity and specificity for pneumococcal vaccine status in adults.^5-8^ Influenza vaccination agreement is higher when self-reporting on the current influenza season compared to the past season suggesting that there may be a difference in immediate and delayed self-reporting of vaccination.^9^ Accurate self-report of other vaccinations, including hepatitis A, may be lower and the accuracy of self-report varies by race/ethnicity and age.^4,10^

Previous research has focused on agreement of vaccine receipt but not on date of vaccine receipt or product. To our knowledge, studies on the accuracy of self-reported COVID-19 vaccination receipt, date, number of doses, or product type are not yet publicly available. Thus, the goal of this analysis was to assess the agreement of self-reported vaccination receipt, number of doses, date(s) received, and vaccine product from two separate surveys (immediate and delayed recall) with vaccination information from participant EHRs.

## METHODS

### Study Sample

The COVID-19 Community Research Partnership (CCRP) is a prospective, multi-site cohort syndromic COVID-19 surveillance study of a convenience sample of adults (18+ years) enrolled from April 2020 through June 2021 primarily through direct email outreach at ten healthcare systems from the mid-Atlantic and southeastern United States (http://www.covid19communitystudy.org/).^11^ Data were collected via a secure, HIPAA-compliant, online platform through March 2022. All participants provided informed consent, and Institutional Review Board (IRB) approval was provided by the Wake Forest School of Medicine IRB. Study sites included: Atrium Health (Charlotte, NC), Campbell University School of Osteopathic Medicine (Lillington, NC), Medstar Health (Columbia, MD), New Hanover Regional Medical Center (Wilmington, NC), Tulane University (New Orleans, LA), University of Maryland – Baltimore (Baltimore, MD), University of Mississippi Medical Center (Jackson, MS), Vidant Health (Greenville, NC), Wake Forest Baptist Health (Winston-Salem, NC), WakeMed Health & Hospitals (Raleigh, NC). For a subset of participants, EHR data were available.

### Ascertainment of Vaccination Date

At baseline, participants self-reported demographic characteristics. COVID-19 symptoms and personal behaviors were self-reported in daily electronic surveillance surveys. Beginning in December 2020, a question was added to the daily electronic surveillance survey asking participants “Have you received a vaccine for COVID-19, since the last survey?” Those responding in the affirmative were asked to enter the vaccination date, the dose number (dose 1 or dose 2), and the vaccine brand (**Appendix**). For the first month of the daily vaccine survey question, the date of receipt was not included; thus, we used the date the survey was completed as a surrogate for date of receipt. Additionally, we developed an algorithm (**Appendix**) to identify data entry errors in the daily survey (e.g., year <2020) and again used the date the survey was entered as the vaccine date in these cases. A secondary supplemental survey was sent to participants in September-October 2021 to ask whether they had received a COVID-19 vaccine, the date of each dose, the dose number (allowing more than two doses) and the vaccine brand. The daily survey was used to evaluate self-reported, immediate recall while the secondary supplemental survey was used for self-reported, delayed recall. In addition, a subset of participants had a record of COVID-19 vaccination in their EHR.

### Participants

Of the 66,403 adult participants enrolled in the CCRP, 46,228 had a data entry on or after December 14, 2020 (first date of COVID-19 vaccine availability in the U.S.). 41,484 adult participants (18+ years) enrolled in the CCRP with COVID-19 vaccination data (receipt, date, and brand) provided on one or more vaccine doses from at least one source (*i.e*., authenticated EHR, immediate recall survey response, or delayed recall survey response) were included in the current analysis (**Appendix Figure 1**). For comparisons of sources, participants with at least two sources of vaccination data (immediate recall self-report, delayed recall self-report, or EHR data) were included in the comparison of vaccination information. Participants missing the date of dose 1 (i.e., only have an entry labeled as dose 2) on the daily survey or with implausible dates/dates with obvious typos (e.g., dates prior to 12/01/2020 for participants not in a clinical trial and dates after the date of entry) on the supplemental survey were excluded.

Included in the primary analysis are 32,122 participants (**Table 1**) with at least two sources of vaccine information for comparison. 29,919 participants are included in the comparison of the two self-reported sources (immediate and delayed recall); 6,628 participants are included in the comparison of immediate recall self-report and EHR vaccine information; and 6,241 participants are included in the comparison of delayed recall self-report and EHR vaccine information (**Table 2**).

**Table 1.** Characteristics of Participants

| **N** | **32,122** |
| --- | --- |
| Age (years) | 54.2 ± 14.5 |
| Sex |  |
| Female | 22,300 (69.4%) |
| Male | 9,819 (30.6%) |
| Unknown | 3 (0.0%) |
| Healthcare Worker Occupation |  |
| No | 24,493 (76.2%) |
| Yes | 7,629 (23.8%) |
| Race/Ethnicity |  |
| American Indian or Alaskan Native | 54 (0.2%) |
| Asian or Pacific Islander | 634 (2.0%) |
| Black or African American | 1,888 (5.9%) |
| Hispanic or Latino | 758 (2.4%) |
| Mixed Ethnicity | 426 (1.3%) |
| Not Specified | 238 (0.7%) |
| White (not Hispanic/Latino) | 28,124 (87.6%) |
| Region |  |
| Deep South | 365 (1.1%) |
| Mid-Atlantic | 11,541 (36.0%) |
| South East | 20,216 (62.9%) |
| County Classification |  |
| Rural | 6,323 (19.7%) |
| Suburban | 7,436 (23.1%) |
| Urban | 18,362 (57.2%) |

**Table 2.** Comparisons of Vaccine Information from Multiple Data Sources (N=32, 122)

|  | **Self-Report** | **Self-Report vs. EHR** |  |
| --- | --- | --- | --- |
|  | **Immediate v Delayed Recall** | **Immediate Recall** | **Delayed Recall** |
| N | 29,919 | 6,628 | 6,241 |
| N Excluded | 1,653 | 567 | 1,072 |
| Reason for Exclusion |  |  |  |
| Likely Booster | 295* | 567<sup>±</sup> | 729<sup>±</sup> |
| Same Date as Entry | 1358<sup>†</sup> | - | 343<sup>†</sup> |
| N for Comparison | 28,266 | 6,061 | 5,169 |
| Number of Doses |  |  |  |
| Match | 26,684 (94.4%) | 5,742 (94.7%) | 5,050 (97.7%) |
| Mismatch | 1,582 (5.6%) | 319 (5.3%) | 119 (2.3%) |
| Dose 1 Date Absolute Difference |  |  |  |
| Median [Q1, Q3] | 0.0 [0.0, 1.0] | 0.0 [0.0, 1.0] | 0.0 [0.0, 1.0] |
| Mean ± SD | 5.7 ± 28.0 | 1.5 ± 10.6 | 2.5 ± 14.8 |
| Dose 1 Date Difference |  |  |  |
| Exact Match | 16,850 (59.6%) | 3,198 (52.8%) | 3,510 (67.9%) |
| +/- 3 Days | 5,812 (20.6%) | 2,679 (44.2%) | 990 (19.2%) |
| +/- 4-7 Days | 2,165 (7.7%) | 39 (0.6%) | 251 (4.9%) |
| +/- 8-14 Days | 1,383 (4.9%) | 31 (0.5%) | 190 (3.7%) |
| +/- 15-30 Days | 1,261 (4.5%) | 87 (1.4%) | 174 (3.4%) |
| +/- >30 Days | 795 (2.8%) | 27 (0.4%) | 54 (1.0%) |
| Product |  |  |  |
| Match | 27,751 (98.2%) | 5,967 (98.4%) | 5,125 (99.1%) |
| Mismatch | 515 (1.8%) | 94 (1.6%) | 44 (0.9%) |

\*Participants were excluded because their only dose in the daily survey (“immediate” recall)
appeared to be their booster dose (e.g., it is after their delayed recall doses 1 and 2, or there is
only one dose recorded in the daily/“immediate” recall survey, and the dose is reported to be
received after October 1, 2021).
<sup>†</sup>Delayed recall survey indicated dose 1 as the same date they took the survey likely due to an
error with the date picker selecting “today’s” date by default. In most cases for these
participants, the dates of doses 1, 2 and 3 were all entered as “today’s” date.
<sup>±</sup>Participants were excluded because their only vaccine date in EHR appeared to be their booster
dose (e.g. does not match their self-reported dose 1 or 2, only one dose in EHR, and only EHR
dose is after October 1, 2021).

### Statistical Analysis

We compared vaccine receipt, number of doses, date of the first dose, and vaccine brand received from self-report (both immediate and delayed recall) to each other and to those available in the EHR. We calculated agreement of receipt, number of doses and vaccine brand. We visually assessed Bland-Altman plots comparing the date of dose 1 (as number of days from 12/14/2020) from the various sources of vaccination dates, representing the mean difference and the 90% confidence interval around the mean difference. Lastly, we compared the presence of vaccine receipt in EHR compared with self-report among all participants with information about receipt and date of at least one dose of a COVID-19 vaccine from at least one source (N=41,488). R version 4.0.13 was used for all statistical analyses.

## RESULTS

### Comparison of Two Sources of Self-Reported Vaccine Information: Immediate vs. Delayed Recall

Prior to comparing the vaccine information, using Bland-Altman plot analysis, we identified 1,653 participants to exclude from further analysis (**Table 2 and Figure 1A**). For most participants, the difference between delayed and immediate report of vaccine information is low, indicated by the fact that most points are included in the confidence interval around zero (green lines). Two point clouds outside the confidence interval at ∼100 days and ∼200 days mean difference in days after 12/14/2020 reflect disagreement, potentially due to participants erroneously entering their booster date as their first dose, or due to entering the day of taking the survey as their date of vaccination. After excluding the flagged participants, 28,266 were available for comparison (**Table 2**). The mean number of days between the initial survey (immediate recall) and supplemental survey (delayed recall) was 205.0 ± 42.6 days (mean ± SD). The number of doses self-reported immediately and self-reported delayed matched for 94.4% (N=26,684). Self-reported immediate recall date of first dose of vaccine was within +/- 7 days of the delayed recall date for 87.9% of participants (**Table 2 and Figure 1B**). As depicted in Figure 1B, a majority of participants were within the 90% confidence interval around the mean difference (green lines), but there are groupings of participants with large differences where one dose may be mislabeled as a second or third dose on a subsequent survey. Self-reported vaccine product (for the first dose recorded) from the delayed recall survey matches the immediate recall survey for 98.2% of participants.

**Figure 1.**
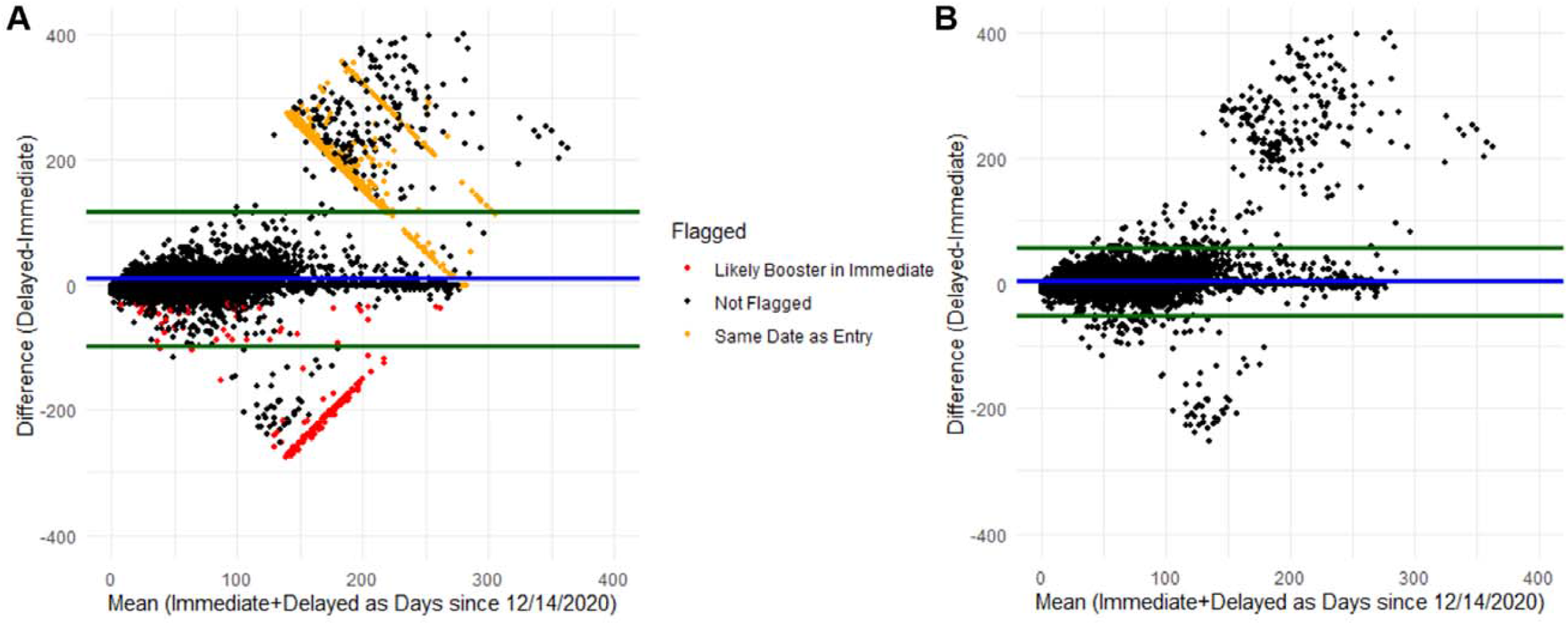
Bland-Altman Plot Comparing Immediate Self-Report and Delayed Self-Report of COVID-19 Vaccination Figure footnote: Blue line represents the mean difference, and green lines represent the 90% confidence interval around the mean difference. Panel A, represents data prior to cleaning identifying likely entry errors. Panel B, represents the data after removing participants with identified errors.

### Comparison of Self-Reported Immediate Recall to EHR Vaccine Information

Prior to comparing the vaccine information, we flagged 567 participants (**Table 2 and Figure 2A)**. After excluding the flagged participants, 6,061 were available for comparison (**Table 2**). The number of doses self-reported and recorded in EHR matched for 94.7%, and among participants who had two doses recorded in their EHR (N=5,826), 96.5% (N=5,621) participants also self-reported two doses. Self-reported date of first dose of vaccine was within +/- 7 days of the EHR date for 97.6% of participants (**Table 2 and Figure 2B**). As depicted in Figure 2B, after excluding the flagged participants, nearly all participants are within the 90% confidence interval around the mean difference (green lines) suggesting that the difference between self-reported date of the first dose and the first date listed in EHR is low. Self-reported vaccine product (for the first dose recorded) from the delayed recall survey matched the EHR product for 98.4% of participants.

**Figure 2.**
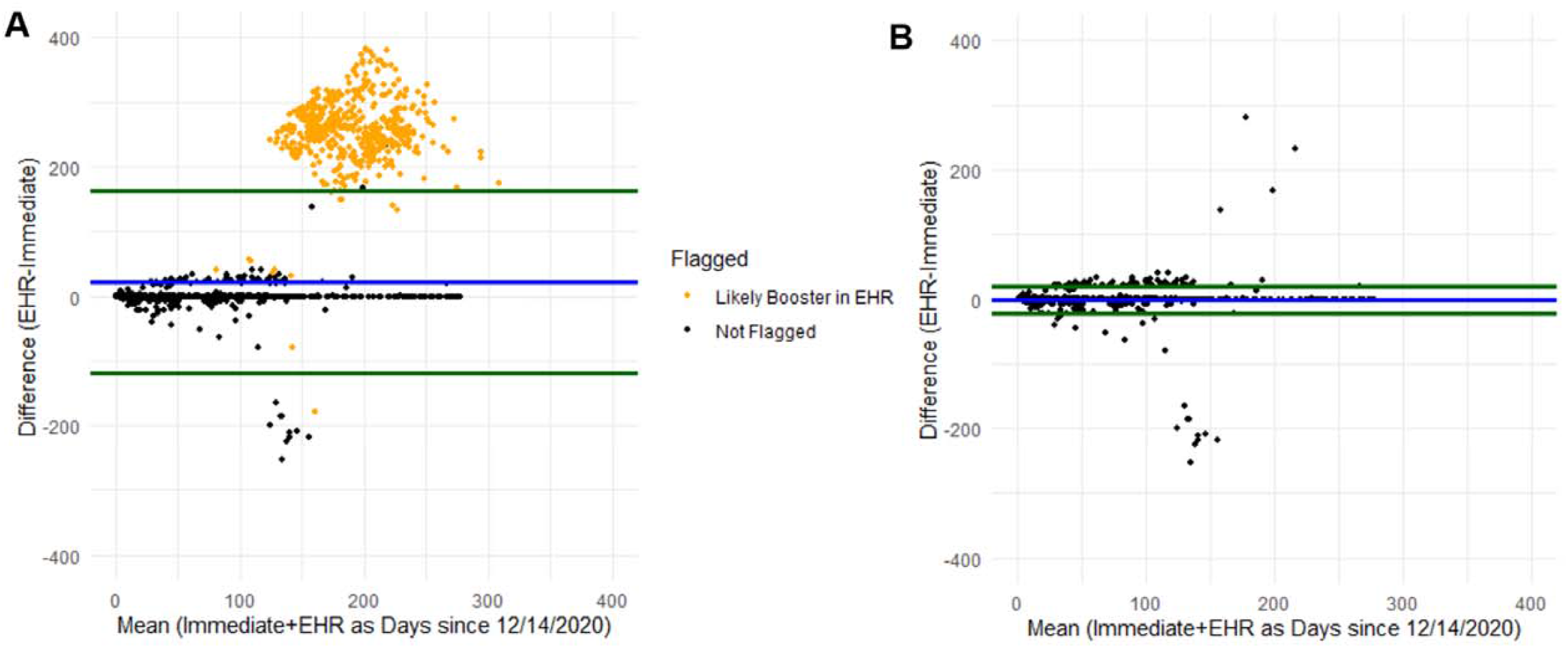
Bland-Altman Plot Comparing Immediate Self-Report and EHR Date of Dose 1 for COVID-19 Vaccination Figure footnote: Blue line represents the mean difference, and green lines represent the 90% confidence interval around the mean difference. Panel A, represents data prior to cleaning identified likely errors. Panel B, represents the data after removing participants with identified errors.

### Comparison of Self-Reported Delayed Recall to EHR Vaccine Information

Prior to comparing the vaccine information, we flagged 1,072 participants (**Table 2 and Figure 3A)**. After excluding the flagged participants, 5,169 were available for comparison (**Table 2**). The number of doses self-reported and recorded in EHR matched for 97.7% of participants and among participants who had two doses recorded in their EHR (N=5,018), 99.8% (N=5,006) participants also self-reported two doses. Self-reported date of first dose of vaccine was within +/- 7 days of the EHR date for 92% of participants (**Table 2 and Figure 3B**). As illustrated in Figure 3B, after excluding the flagged participants, the majority of participants are within the 90% confidence interval around the mean difference (green lines) with a subset of participants in the top-center that may have mislabeled doses and participants in the bottom-center that have dates similar to the date the supplemental survey was released. Self-reported vaccine product (for the first dose recorded) from the delayed recall survey matched the EHR product for 99.1% of participants.

**Figure 3.**
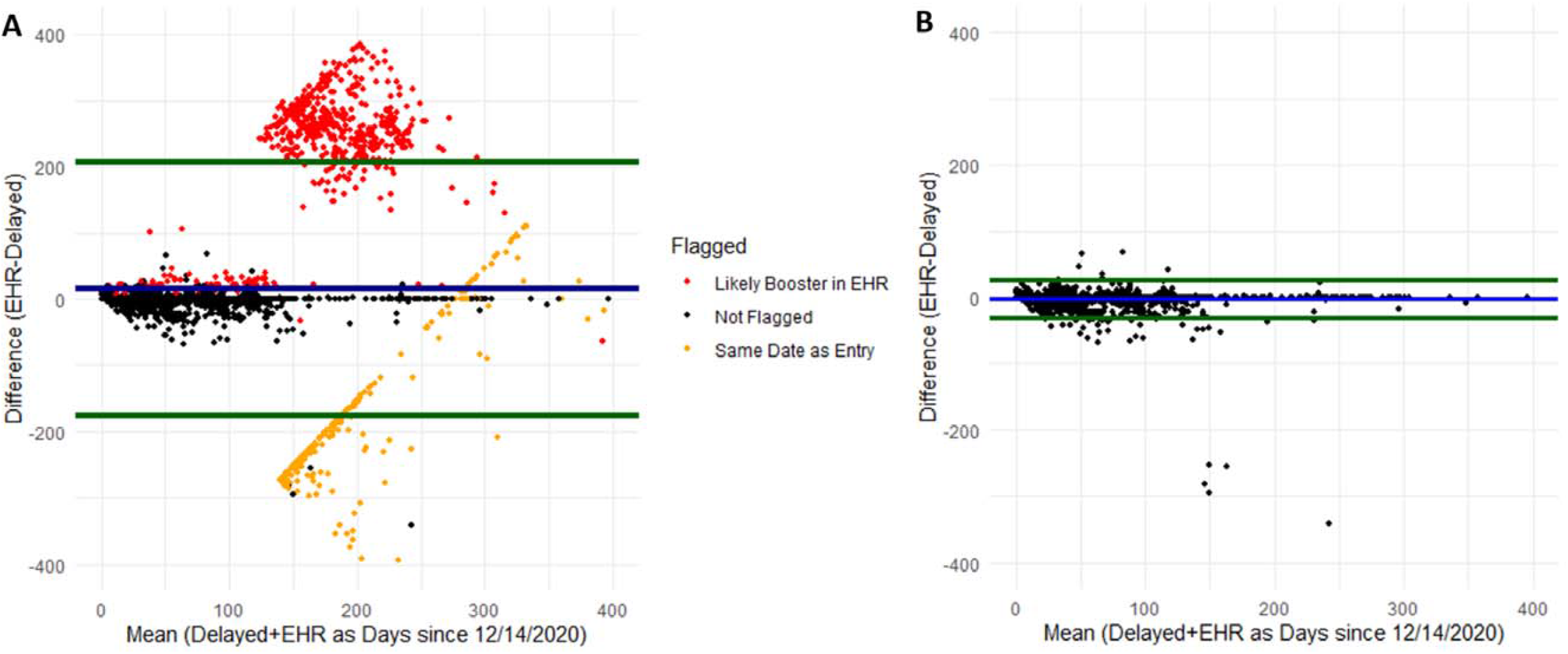
Bland-Altman Plot Comparing Delayed Self-Report and EHR Date of Dose 1 for COVID-19 Vaccination Figure footnote: Blue line represents the mean difference and green lines represent the 90% confidence interval around the mean difference. Panel A represents data prior to cleaning identified likely errors. Panel B represents the data after removing participants with identified errors.

### Comparison of Vaccine Receipt

As a secondary analysis, we compared the presence of vaccine receipt in EHR compared with self-report among all participants with information about receipt and date of at least one dose of a COVID-19 vaccine from at least one source (N=41,484). Among participants with at least one survey entry after March 1, 2021 when vaccines were widely available (N=39,735), 19.6% (N=7,786) participants had evidence of vaccination in their EHR, and 94.9% of them self-reported vaccination in one of the two surveys. Among participants with at least one survey entry after the supplemental survey was sent on September 16 (N=32,103), participants 19.6% (N=6,303) had evidence of vaccination in their EHR, and 99% of them self-reported vaccination in the supplemental survey (**Appendix Table 1**).

## DISCUSSION

In the CCRP cohort, we found strong agreement between self-reported and EHR information pertaining to receipt of the SARS-CoV-2 vaccinations including number of doses, date of receipt of the first dose, and product details. In our cohort, there was high agreement between immediate and delayed recall self-reported vaccination information in terms of number of doses reported (94% agreement), vaccine product (98% agreement), and date of first vaccine dose (88% within +/- 7 days). While number of doses and vaccine product had similar agreement with EHR vaccination information for the immediate and delayed recall self-report, the immediate recall date of dose 1 was more closely aligned with the date found in EHR, with 98% of participants within +/-7 days for immediate recall compared to 92% for delayed recall. This difference is likely explained by the timing between vaccination and report. On average, 205 days passed between the daily survey (immediate recall) and supplemental survey (delayed recall). While the coverage of SARS-CoV-2 vaccination data in participant electronic medical records was only available for about a fifth of participants with linked EHR, nearly all of them (95%) also self-reported vaccination. Among participants with data in two sources, comparison of the details (number of doses, dates, and product) revealed the strength of self-reported COVID-19 vaccination data.

Our results contribute evidence about the use of self-reported COVID-19 vaccination dates. To our knowledge, this is the first study to evaluate the accuracy of self-reported COVID-19 vaccination information. Researchers have shown that in test-negative studies of VE, self-reported influenza vaccination results in biased estimates, suggesting the importance of validating self-reported dates.^12,13^ However, they also found that the greatest impact for influenza VE estimation occurred when vaccination coverage was low. About half of Americans typically receive an influenza vaccine annually^14^ compared with over 80% of Americans having received at least one dose of a SARS-CoV-2 vaccine.^15^ This difference in vaccine coverage (50% for influenza vs. 80% for COVID-19) taken together with our findings of high agreement of self-report and EHR vaccine information suggests that the use of self-reported SARS-CoV-2 vaccination may have less of an impact on biasing VE estimates than observed with influenza.

While EHR vaccination information may be considered the gold standard, we note that the lack of documentation of vaccination in EHR likely does not indicate that no vaccination had been obtained. In the CCRP, only a subset of our participants had documentation of SARS-CoV-2 vaccination in their medical records. Moreover, vaccination dates in EHR do not guarantee delivery of the vaccine by the healthcare system and may be data entry from a patient’s vaccination card or patient recall during subsequent routine medical visits. Data entry errors are also possible in medical records.

The COVID-19 vaccines are unique from other vaccinations (e.g. influenza) in their distribution at largely mass vaccination sites and pharmacies (e.g. CVS, Walgreens) rather than from one’s primary care provider or employer.^16,17^ The rollout of the vaccines in December 2020 and early 2021 was a memorable milestone in the pandemic which may have made the date of receipt of first dose more memorable than an annual influenza vaccine. Additionally, the vaccine brands were widely discussed by the media, making them part of popular culture and easily recognizable.^18^

The CCRP comparison of self-reported and EHR vaccination information had several strengths including the large number of participants with self-reported and EHR data available for comparison. Many participants self-reported their vaccine data twice: once in real time as vaccines were being administered and once several months later for confirmation. Our study has a number of important limitations. The CCRP is a U.S. healthcare system-based convenience sample that is predominantly non-Hispanic White and about a quarter are healthcare workers.

Participants active in the study are likely to be generally interested in their health and may have greater awareness of the details surrounding vaccination. These results may not be consistent in populations of varying race/ethnicity, occupation, education level, health literacy, or region. Importantly, the results are relevant only to the U.S. population where the details of COVID-19 vaccines have been discussed extensively in the media and may not be generalizable to other regions with different media landscapes, and vaccine attitudes and availability. Additionally, there were significant differences between participants with and without documentation of vaccination in EHR: female participants, healthcare workers, participants in the South East, and participants residing in urban areas were more likely to have vaccine information in their EHR (**Appendix Table 2**). There were also differences by enrollment site. This difference in availability of EHR data highlights the importance of accepting self-reported vaccine dates particularly where EHR vaccination data may be sparse. Thus, we have not computed sensitivity or specificity of self-reported receipt of vaccination. Additionally, due to the structure of the vaccination data we received from EHR (unnumbered doses), it was difficult to identify the dose number especially when one or more doses were likely missing. The self-reported dates for CCRP were also flawed. Initially, we did not ask the date of receipt for vaccination, and later the data entry system allowed the date field to be left blank. Insufficient data checks were in place to prevent the entry of nonsensical dates (e.g. typos, multiple doses labeled dose 1, dose 2 occurring before dose 1, etc.). For the immediate recall survey, we used the date the daily survey was entered as a surrogate for the date of receipt. For the supplemental survey (delayed recall), the date picker that allowed users to enter a date either through text input, or by choosing a date from the calendar defaulted to “today’s” date which led to issues with data quality. For the CCRP analyses, we took advantage of having three sources of vaccination data and compared the three to identify the dates most likely to be correct for each participant taking into account agreement between more than one source, timing between doses, and timing of entry of immediate recall survey.

## CONCLUSIONS

Our findings suggest that self-reported dates and product details can be an effective surrogate when medical records are unavailable in large observational studies. There are limitations to using medical records for COVID-19 vaccination as they do not always note if vaccination actually occurred; thus, self-reported dates may provide us with more data and may be the simplest option for obtaining vaccination status and details. It is important to have robust data entry rules in place to avoid errors at the point of entry and when possible, to collect data in real time about vaccine receipt as the self-reported data may be of higher quality than delayed recall. A secondary confirmation of vaccine data on a subset of participants using EHR will provide evidence of validity.

## Data Availability

Results of the COVID-19 CRP are being disseminated on the study website (https://www.covid19communitystudy.org/) as well as in publications and presentations in medical journals and at scientific meetings. At end of the study, the databases will be made publicly available in a de-identified manner according to CDC and applicable U.S. Federal policies.

## ACKNOWLEDGMENTS

The COVID-19 Community Research Partnership gratefully acknowledges the commitment and dedication of the study participants. Programmatic, laboratory, and technical support was provided by Vysnova Partners, Inc., Javara, Inc., Oracle Corporation, LabCorp, Scanwell Health, and Neoteryx. This publication was supported by the Centers for Disease Control and Prevention (CDC) [contract #75D30120C08405] and the CARES Act of the U.S. Department of Health and Human Services (HHS) [Contract # NC DHHS GTS #49927]. Fifty percent of the current project was funded by the CDC/HHS award and fifty percent by the CARES Act/HHS award. The Partnership is listed in clinicaltrials.gov (NCT04342884). The findings and conclusions in this report are those of the authors and do not necessarily represent the official position of the Centers for Disease Control and Prevention, HHS, or the U.S. Government. A complete list of Study Sites, investigators, and staff can be found in the Appendix.

## APPENDIX

**Appendix Table 1.** Comparing Vaccine Self-Reported Vaccine Receipt by EHR Vaccine Receipt (among participants with information about receipt and date of at least one dose of a COVID-19 vaccine from at least one source, N=41,488)

| *Among participants with at least one survey entry after 03/01/2021 (N=39,735)* |  |  |
| --- | --- | --- |
|  | **EHR +** | **EHR -** |
| N | 7,786 | 31,949 |
| Immediate Recall |  |  |
| Self-Report + | 6,479 (83.2%) | 31,302 (98.0%) |
| Self-Report - | 1,307 (16.8%) | 647 (2.0%) |
| Delayed Recall |  |  |
| Self-Report + | 6,236 (80.1%) | 25,171 (78.8%) |
| Self-Report - | 1,550 (19.9%) | 6,778 (21.2%) |
| Either Self-Report |  |  |
| Self-Report + | 7,386 (94.9%) | 31,949 (100.0%) |
| Self-Report - | 400 (5.1%) | 0 (0.0%) |
| *Among participants with at least one survey entry after 09/16/2021 (N=32,064)* |  |  |
|  | **EHR +** | **EHR -** |
| N | 6,303 | 25,800 |
| Delayed Recall |  |  |
| Self-Report + | 6,240 (99.0%) | 25,214 (97.7%) |
| Self-Report - | 63 (1.0%) | 586 (2.3%) |

**Appendix Table 2.** Comparing Characteristics of Participants with Vaccine Information in EHR (among participants with information about receipt and date of at least one dose of a COVID-19 vaccine in at least one source, N=41,484)

|  | Without Vaccine in EHR<br>N=32,972 | With Vaccine in EHR<br>N=8,512 |
| --- | --- | --- |
| Site |  |  |
| Atrium | 2,689 (8.2%) | 4,232 (49.7%) |
| Campbell | 447 (1.4%) | 0 (0.0%) |
| Maryland | 4,683 (14.2%) | 0 (0.0%) |
| MedStar | 8,551 (25.9%) | 1,292 (15.2%) |
| Mississippi | 71 (0.2%) | 272 (3.2%) |
| New Hanover | 369 (1.1%) | 334 (3.9%) |
| Tulane | 166 (0.5%) | 0 (0.0%) |
| Vidant | 752 (2.3%) | 0 (0.0%) |
| Wake Forest | 13,404 (40.6%) | 1,495 (17.6%) |
| Wake Med | 1,842 (5.6%) | 887 (10.4%) |
| Age (years) | 53.2 ± 14.8 | 51.4 ± 15.1 |
| Sex |  |  |
| Female | 22,582 (68.5%) | 6,179 (72.6%) |
| Male | 10,385 (31.5%) | 2,333 (27.4%) |
| Unspecified | 5 (0.0%) | 0 (0.0%) |
| Healthcare Worker |  |  |
| No | 27,001 (81.9%) | 4,433 (52.1%) |
| Yes | 5,971 (18.1%) | 4,079 (47.9%) |
| Race/Ethnicity |  |  |
| Am Ind/Alaskan Native | 98 (0.3%) | 11 (0.1%) |
| Asian/Pac Isl | 647 (2.0%) | 244 (2.9%) |
| Black/African Am | 2,004 (6.1%) | 555 (6.5%) |
| Hispanic/Latino | 834 (2.5%) | 218 (2.6%) |
| Mixed Ethnicity | 445 (1.3%) | 132 (1.6%) |
| Not Specified | 308 (0.9%) | 62 (0.7%) |
| Non-Hispanic White | 28,636 (86.8%) | 7,290 (85.6%) |
| Region of Enrollment |  |  |
| Deep South | 237 (0.7%) | 272 (3.2%) |
| North Atlantic | 13,234 (40.1%) | 1,292 (15.2%) |
| South East | 19,501 (59.1%) | 6,948 (81.6%) |
| County of Residence |  |  |
| Rural | 7,218 (21.9%) | 1,072 (12.6%) |
| Suburban | 7,338 (22.2%) | 2,093 (24.6%) |
| Urban | 18,415 (55.9%) | 5,347 (62.8%) |

$P < 0.001$ for all comparisons. P-values for Pearson's Chi-squared test for categorical variables and Welch's two-sample t-test for continuous variables.

**Appendix Figure 1.**
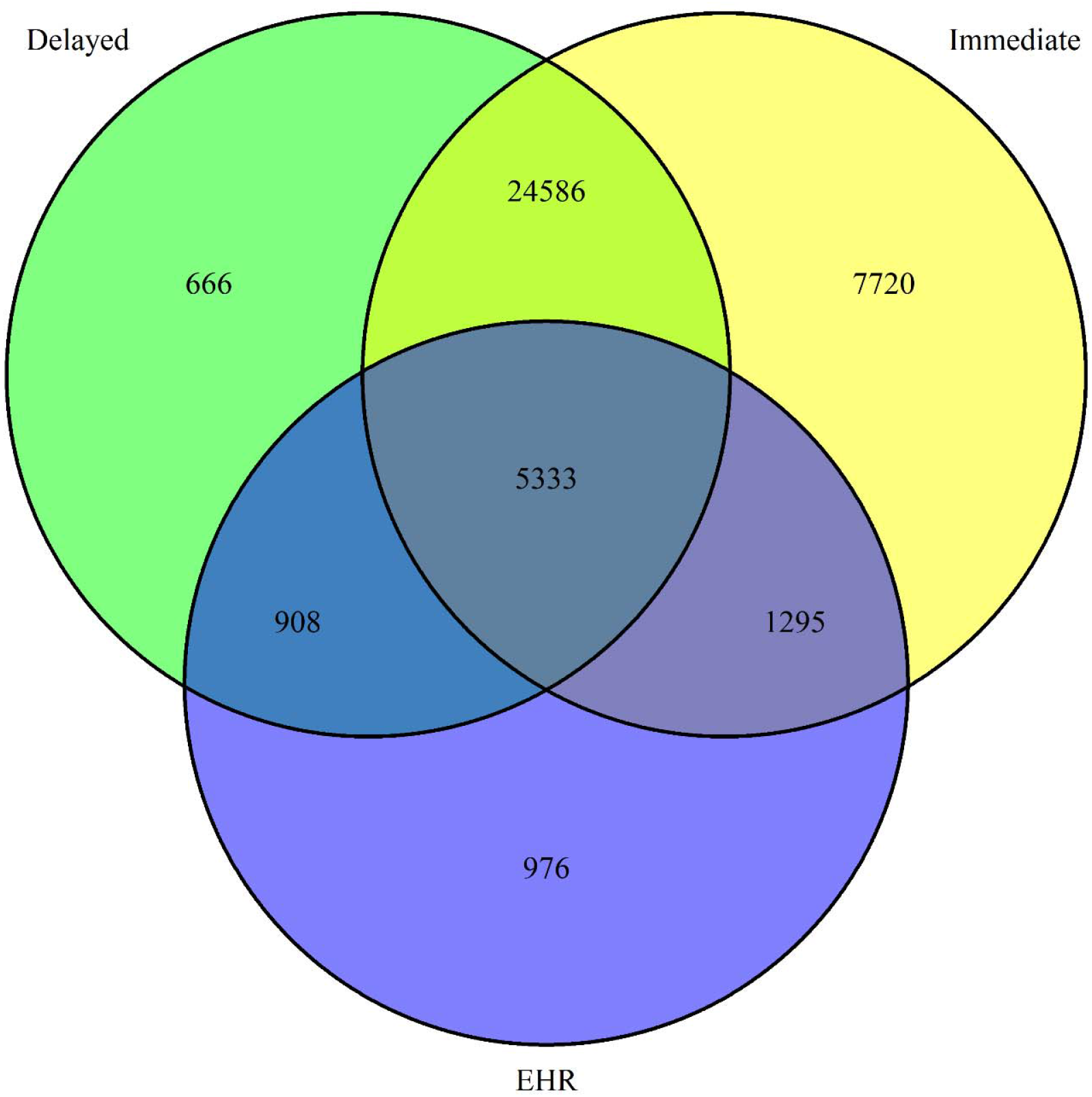
Venn Diagram Depicting Number of Participants with COVID-19 Vaccine Information Available from each Source

### APPENDIX A. Vaccine Questions

##### Daily Questionnaire (Immediate Recall) for Adult participants

Please answer the following questions based on what you are experiencing between your last update and now.

- Have you received a vaccine for COVID-19, since your last update?
  - Yes
  - No
  - If YES:
    ▪ Was this part of a clinical trial?
      - Yes
      - No
      - I don’t know
    ▪ Which vaccine:
      - I do not know
      - AstraZeneca - AZD1222
      - Johnson & Johnson - Ad26.COV2.S
      - Moderna - mRNA
      - Pfizer - BNT162
      - Other
    ▪ Which dose:
      - Dose 1
      - Dose 2
    ▪ Vaccination date? (If Known)

##### Supplemental Questionnaire (Delayed Recall) for Adult participants

**Questions added to daily survey to replace existing questions:**

1. How many doses of the COVID-19 Vaccine have you received in total? *
  - Zero
  - One
  - Two
  - Three
  - Four or more
    - Please enter total number of COVID-19 vaccine doses you have received. [text box]

***If a person chooses this is my first dose, two, three, or four or more ask question***

2. Tell us about your **first** COVID-19 vaccine dose. Was this part of a clinical trial? **(If Yes, end of questions)**
  - Yes
  - No
  - I don’t know
3. What was your first dose of the COVID-19 Vaccine?*
  - Moderna
  - Janssen/Johnson & Johnson
  - Pfizer-BioNTech
  - Other/ I don’t know
4. When did you receive your first COVID-19 Vaccine?
  - Calendar Function

***If a person chose two, three, or four or more in Q1 ask question***

5. Tell us about your **second** COVID-19 vaccine dose. What was your second dose of the COVID-19 Vaccine?*
  - Moderna
  - Janssen/Johnson & Johnson
  - Pfizer-BioNTech
  - Other/ I don’t know
6. When did you receive your second COVID-19 Vaccine?
  - Calendar Function

***If a participant selected ”three or four or more” on question 1***

7. Tell us about your **third** COVID-19 vaccine dose. What was your second dose of the COVID-19 Vaccine?*
  - Moderna
  - Janssen/Johnson & Johnson
  - Pfizer-BioNTech
  - Other/ I don’t know
8. When did you receive your third COVID-19 Vaccine?
  - Calendar Function
9. Why did you seek an additional dose of COVID-19 Vaccine [select all that applies]
  - Additional protection
  - Immunocompromised
  - Greater than 8-months since I received my vaccine
  - Other
    - Text Box for additional response

***If a participant selected ”one or two” on question 1 on last survey***

10. Since your last survey, have you had an additional COVID-19 Vaccine dose?*
  - Yes
  - No
11. What was your [dose number] of the COVID-19 Vaccine?*
  - Moderna
  - Janssen/Johnson & Johnson
  - Pfizer-BioNTech
  - Other/ I don’t know
12. When did you receive your COVID-19 Vaccine
  - Calendar Function
13. Why did you seek an additional dose of COVID-19 Vaccine [select all that applies]
  - Additional protection
  - Immunocompromised
  - Greater than 8-months since I received my vaccine
  - Other
    - Text Box for additional response

Once participant answers question 1, question 1-9 should go away. Going forward, participant should be asked Question 10.

We should bring up the next set of vaccine questions (i.e. vaccine 1, 2, or 3) based on the what the participant last completed.

After someone with J&J had at least 2 doses and someone with Moderna/Pfizer had 3 doses, question series goes away.

### Authorship Appendix: The COVID-19 Research Group (*Site Principal Investigator)

**Wake Forest School of Medicine:** Thomas F Wierzba PhD, MPH, MS*, John Walton Sanders, MD, MPH, David Herrington, MD, MHS, Mark A. Espeland, PhD, MA, John Williamson, PharmD, Morgana Mongraw-Chaffin, PhD, MPH, Alain Bertoni, MD, MPH, Martha A. Alexander-Miller, PhD, Paola Castri, MD, PhD, Allison Mathews, PhD, MA, Iqra Munawar, MS, Austin Lyles Seals, MS, Brian Ostasiewski, Christine Ann Pittman Ballard, MPH, Metin Gurcan, PhD, MS, Alexander Ivanov, MD, Giselle Melendez Zapata, MD, Marlena Westcott, PhD, Karen Blinson, Laura Blinson, Mark Mistysyn, Donna Davis, Lynda Doomy, Perrin Henderson, MS, Alicia Jessup, Kimberly Lane, Beverly Levine, PhD, Jessica McCanless, MS, Sharon McDaniel, Kathryn Melius, MS, Christine O’Neill, Angelina Pack, RN, Ritu Rathee, RN, Scott Rushing, Jennifer Sheets, Sandra Soots, RN, Michele Wall, Samantha Wheeler, John White, Lisa Wilkerson, Rebekah Wilson, Kenneth Wilson, Deb Burcombe, Georgia Saylor, Megan Lunn, Karina Ordonez, Ashley O’Steen, MS, Leigh Wagner.

**Atrium Health:** Michael S. Runyon MD, MPH*, Lewis H. McCurdy MD*, Michael A. Gibbs, MD, Yhenneko J. Taylor, PhD, Lydia Calamari, MD, Hazel Tapp, PhD, Amina Ahmed, MD, Michael Brennan, DDS, Lindsay Munn, PhD RN, Keerti L. Dantuluri, MD, Timothy Hetherington, MS, Lauren C. Lu, Connell Dunn, Melanie Hogg, MS, CCRA, Andrea Price, Marina Leonidas, Melinda Manning, Whitney Rossman, MS, Frank X. Gohs, MS, Anna Harris, MPH, Jennifer S. Priem, PhD, MA, Pilar Tochiki, Nicole Wellinsky, Crystal Silva, Tom Ludden PhD, Jackeline Hernandez, MD, Kennisha Spencer, Laura McAlister.

**MedStar Health Research Institute:** William Weintraub MD*, Kristen Miller, DrPH, CPPS*, Chris Washington, Allison Moses, Sarahfaye Dolman, Julissa Zelaya-Portillo, John Erkus, Joseph Blumenthal, Ronald E. Romero Barrientos, Sonita Bennett, Shrenik Shah, Shrey Mathur, Christian Boxley, Paul Kolm, PhD, Ella Franklin, Naheed Ahmed, Moira Larsen.

**Tulane:** Richard Oberhelman MD*, Joseph Keating PhD*, Patricia Kissinger, PhD, John Schieffelin, MD, Joshua Yukich, PhD, Andrew Beron, MPH, Johanna Teigen, MPH.

**University of Maryland School of Medicine:** Karen Kotloff MD*, Wilbur H. Chen MD, MS*, DeAnna Friedman-Klabanoff, MD, Andrea A. Berry, MD, Helen Powell, PhD, Lynnee Roane, MS, RN, Reva Datar, MPH, Colleen Reilly.

**University of Mississippi:** Adolfo Correa MD, PhD*, Bhagyashri Navalkele, MD, Yuan-I Min, PhD, Alexandra Castillo, MPH, Lori Ward, PhD, MS, Robert P. Santos, MD, MSCS, Pramod Anugu, Yan Gao, MPH, Jason Green, Ramona Sandlin, RHIA, Donald Moore, MS, Lemichal Drake, Dorothy Horton, RN, Kendra L. Johnson, MPH, Michael Stover.

**Wake Med Health and Hospitals:** William H. Lagarde MD*, LaMonica Daniel, BSCR.

**New Hanover:** Patrick D. Maguire MD*, Charin L. Hanlon, MD, Lynette McFayden, MSN, CCRP, Isaura Rigo, MD, Kelli Hines, BS, Lindsay Smith, BA, Monique Harris, CCRP, Belinda Lissor, AAS, CCRP, Vivian Cook, MA, MPH, Maddy Eversole, BS, Terry Herrin, BS, Dennis Murphy, RN, Lauren Kinney, BS, Polly Diehl, MS, RHIA, Nicholas Abromitis, BS, Tina St. Pierre, BS, Bill Heckman, Denise Evans, Julian March, BA, Ben Whitlock, CPA, MSA, Wendy Moore, BS, AAS, Sarah Arthur, MSW, LCSW, Joseph Conway.

**Vidant Health:** Thomas R. Gallaher MD*, Mathew Johanson, MHA, CHFP, Sawyer Brown, MHA, Tina Dixon, MPA, Martha Reavis, Shakira Henderson, PhD, DNP, MS, MPH, Michael Zimmer, PhD, Danielle Oliver, Kasheta Jackson, DNP, RN, Monica Menon, MHA, Brandon Bishop, MHA, Rachel Roeth, MHA.

**Campbell University School of Osteopathic Medicine:** Robin King-Thiele DO*, Terri S. Hamrick PhD*, Abdalla Ihmeidan, MHA, Amy Hinkelman, PhD, Chika Okafor, MD (Cape Fear Valley Medical Center), Regina B. Bray Brown, MD, Amber Brewster, MD, Danius Bouyi, DO, Katrina Lamont, MD, Kazumi Yoshinaga, DO, (Harnett Health System), Poornima Vinod, MD, A. Suman Peela, MD, Giera Denbel, MD, Jason Lo, MD, Mariam Mayet-Khan, DO, Akash Mittal, DO, Reena Motwani, MD, Mohamed Raafat, MD (Southeastern Health System), Evan Schultz, DO, Aderson Joseph, MD, Aalok Parkeh, DO, Dhara Patel, MD, Babar Afridi, DO (Cumberland County Hospital System, Cape Fear Valley).

**George Washington University Data Coordinating Center:** Diane Uschner PhD*, Sharon L. Edelstein, ScM, Michele Santacatterina, PhD, Greg Strylewicz, PhD, Brian Burke, MS, Mihili Gunaratne, MPH, Meghan Turney, MA, Shirley Qin Zhou, MS, Ashley H Tjaden, MPH, Lida Fette, MS, Asare Buahin, Matthew Bott, Sophia Graziani, Ashvi Soni, MS, Guoqing Diao, PhD, Jone Renteria, MS.

**George Washington University Mores Lab:** Christopher Mores, PhD, Abigail Porzucek, MS. Oracle Corporation: Rebecca Laborde, Pranav Acharya. Sneez LLC: Lucy Guill, MBA, Danielle Lamphier, MBA, Anna Schaefer, MSM, William M. Satterwhite, JD, MD. Vysnova Partners: Anne McKeague, PhD, Johnathan Ward, MS, Diana P. Naranjo, MA, Nana Darko, MPH, Kimberly Castellon, BS, Ryan Brink, MSCM, Haris Shehzad, MS, Derek Kuprianov, Douglas McGlasson, MBA, Devin Hayes, BS, Sierra Edwards, MS, Stephane Daphnis, MBA, Britnee Todd, BS. Javara Inc: Atira Goodwin.

**External Advisory Council:** Ruth Berkelman, MD, Emory, Kimberly Hanson, MD, U of Utah, Scott Zeger, PhD, Johns Hopkins, Cavan Reilly, PhD, U. of Minnesota, Kathy Edwards, MD, Vanderbilt, Helene Gayle, MD MPH, Chicago Community Trust, Stephen Redd.

